# Serosurveillance of dengue infection and correlation with mosquito pools for dengue virus positivity during the COVID-19 pandemic in Tamil Nadu, India – A state-wide cross-sectional cluster randomized community-based study

**DOI:** 10.1101/2024.06.07.24308595

**Authors:** Sivaprakasam T. Selvavinayagam, Sathish Sankar, Yean K. Yong, Abdul R. Anshad, Samudi Chandramathi, Anavarathan Somasundaram, Sampath Palani, Parthipan Kumarasamy, Roshini Azhaguvel, Ajith B. Kumar, Sudharshini Subramaniam, Manickam Malathi, Venkatachalam Vijayalakshmi, Manivannan Rajeshkumar, Anandhazhvar Kumaresan, Ramendra P. Pandey, Nagarajan Muruganandam, Natarajan Gopalan, Meganathan Kannan, Amudhan Murugesan, Pachamuthu Balakrishnan, Siddappa N. Byrareddy, Aditya P. Dash, Marie Larsson, Vijayakumar Velu, Esaki M. Shankar, Sivadoss Raju

**Affiliations:** State Public Health Laboratory, Directorate of Public Health and Preventive Medicine, DMS Campus, Teynampet 600 018, Chennai, Tamil Nadu, India; Centre for Infectious Diseases, Department of Microbiology, Saveetha Dental College and Hospitals, Saveetha Institute of Medical and Technical Sciences, Saveetha University, Chennai 600077, Tamil Nadu, India; Laboratory Centre, Xiamen University Malaysia, 43900 Sepang, Selangor, Malaysia; Infection and Inflammation, Department of Biotechnology, Central University of Tamil Nadu, Thiruvarur 610 005, India; Department of Medical Microbiology, Faculty of Medicine, University of Malaya, Lembah Pantai, Kuala Lumpur, Malaysia; Institute of Community Medicine, Madras Medical College, Chennai, Tamil Nadu, India; Institute of Vector Control and Zoonoses, Hosur, 635126, Tamil Nadu; School of Health Sciences and Technology, UPES, Dehradun, 248007, Uttarakhand, India; Regional Medical Research Centre, Indian Council of Medical Research, Port Blair, Andaman and Nicobar Islands, India; Department of Epidemiology and Public Health, Central University of Tamil Nadu, Thiruvarur 610 005, India; Blood and Vascular Biology, Department of Biotechnology, Central University of Tamil Nadu, Thiruvarur 610 005, India; Department of Microbiology, Government Theni Medical College and Hospital, Theni, Tamil Nadu, India; Center for Infectious Diseases, Saveetha Medical College and Hospital, Saveetha Institute of Medical and Technical Sciences, Saveetha University, Chennai, Tamil Nadu, India; Department of Pharmacology and Experimental Neuroscience, University of Nebraska Medical Center, Omaha, NE 68131, USA; Asian Institute of Public Health University, Bhubaneswar, Odisha, India; Division of Molecular Medicine and Virology, Department of Biomedical and Clinical Sciences, Linköping University, 58 185 Linköping, Sweden; Department of Pathology and Laboratory Medicine, Emory University School of Medicine, Division of Microbiology and Immunology, Emory National Primate Research Center, Emory Vaccine Center, Atlanta, GA, 30329, USA

**Keywords:** COVID-19, Dengue, Serosurveillance, Vector-borne disease

## Abstract

**Background:** Dengue is a vector-borne viral disease impacting millions across the globe. Nevertheless, akin to many other diseases, reports indicated a decline in dengue incidence and seroprevalence during the COVID-19 pandemic (2020-22). This presumably could be attributed to reduced treatment-seeking rates, under-reporting, misdiagnosis, disrupted health services and reduced exposure to vectors due to lockdowns. Scientific evidence on dengue virus (DENV) disease during the COVID-19 pandemic is limited globally.

**Methods:** A cross-sectional, randomized cluster sampling community-based survey was carried out to assess anti-dengue IgM and IgG and SARS-CoV-2 IgG seroprevalence across all 38 districts of Tamil Nadu, India. The prevalence of DENV in the Aedes mosquito pools during 2021 was analyzed and compared with previous and following years of vector surveillance for DENV by real-time PCR.

**Findings:** Results implicate that both DENV-IgM and IgG seroprevalence and mosquito viral positivity were reduced across all the districts. A total of 13464 mosquito pools and 5577 human serum samples from 186 clusters were collected. Of these, 3·76% of mosquito pools were positive for DENV. In the human sera, 4·12% were positive for DENV IgM and 6·4% were positive for DENV IgG. The anti-SARS-CoV-2 antibody titres correlated with dengue seropositivity with a significant association whereas vaccination status significantly correlated with dengue IgM levels.

**Interpretation:** Continuous monitoring of DENV seroprevalence, especially with the evolving variants of the SARS-CoV-2 virus and surge in COVID-19 cases will shed light on the transmission and therapeutic attributes of dengue infection.

## Introduction

Dengue represents a global arboviral public health threat, and is caused by four serotypes of dengue virus (DENV1-4). *Aedes* mosquitoes (*Ae. aegypti* and *Ae. albopictus*), act as vectors that dwell in the tropical and subtropical world making the disease hyperendemic across Asia and South America, Africa, the Middle East^1,2^ and other temperate parts of the world^3^. The single-stranded positive-sense RNA-laden Flavivirus causes frequent concurrent epidemics involving different serotypes. While DENV2 appears to be associated with severe disease, there is evidence of distribution of all DENV serotypes in Asia^4^. Dengue is classified as primary and secondary based on IgM:IgG ratio, and two types, viz. dengue without warning signs (DWWS) and dengue with warning signs (DWS) based on clinical manifestations^5,6^. The prognosis of dengue is determined by antibody-dependent enhancement (ADE), viral dynamics, and pre-existing antibody titers^7^. However, protean clinical manifestations, serotype heterogeneity, and co-infections pose a substantial challenge to patient management.

There is a growing interest in prevailing infections post-SARS-CoV-2 pandemic. As with other infections, there has been a shift in the trend of dengue in 2020-22, when COVID-19 was taking a toll. Several studies reported a 16-97% decrease in dengue cases during the pandemic^8–10^. There have been reports of concomitant dengue disease together with other infectious agents, including SARS-CoV-2^11–14^. The pressure that prevailed on COVID-19 pandemic raised concerns over the lack of attention to dengue diagnosis, reduced treatment-seeking rates, potential for misdiagnosis, reduced availability of laboratory testing for dengue, and negative impact of lockdowns^10^. There has been a declining trend in dengue post-COVID-19 following an upsurge in 2019^15^, likely due to global imposition of lockdowns^16^. In India, dengue incidence was reported to be ∼188,000 (2017), 101,192 (2018) and 157,315 (2019) cases. However, the frequency of dengue declined abruptly to 45,585 (71%) (https://ncvbdc.mohfw.gov.in) in 2020^17^.

Studies reporting dengue decline during the pandemic were often based on serological investigations (NS1/IgM/IgG). Our state-wide entomological surveillance and vector control data indicated a significant reduction of DENV-positive mosquito pools in 2020 that remained low until 2023. This further substantiated our assumption of reduced DENV transmission due to lockdowns. We hypothesized that there is a correlation trend between the SARS-CoV-2 IgG as well as anti-DENV IgM and IgG titers. Possibly, antibodies to SARS-CoV-2 could hinder the circulation of DENV either by protective cross-reaction, antigenic similarity or by masked effects of ADE^18^. The cross-reactive nature of anti-SARS-CoV-2 was reported against various antigens and vaccines^19^. Antibodies to spike and receptor-binding domain (S1-RBD) have been shown to cross-react with both DENV envelope protein (E) and non-structural protein 1 (NS1) in experimental animals^20^.

Constant monitoring of disease prevalence and entomological surveillance together with risk factors of viral transmission are critical for highly endemic countries like India. Here, we conducted a community-based, cross-sectional, cluster randomized survey to assess the seroprevalence of dengue and DENV positivity in aedine mosquito vectors in Tamil Nadu, India in December 2021. The primary and secondary DENV infections along with the antibody titres were correlated with the SARS-CoV-2 IgG in the population.

## Methods

### Mosquito sampling

The eggs, larvae, and adult Aedes mosquitoes were collected from across all the 38 districts of Tamil Nadu from indoors and outdoors. The sampling and testing are being carried out as part of the routine surveillance program since 2016 for the prevention and control of vector-borne diseases by the Department of Public Health, Tamil Nadu, India. Here, we compared and analyzed the samples collected during 2016 to mid-2024 for possible correlation with the seroprevalence of SARS-CoV-2 and DENV. The adult female Aedes mosquitoes captured were identified and isolated using a standard method^8^, and were transported to the processing laboratory. The eggs hatched after an incubation period of 15 days at the Regional Entomology Laboratory. The larvae and adults were identified and the dried adult mosquito samples were transported in zip-lock covers or microcentrifuge tubes to the State Public Health Laboratory (SPHL), Chennai, and the Institute of Vector Control and Zoonosis (IVCZ), Hosur, India.

### Sample processing

Engorged adult female mosquito pools (n=25) collected from specific trap areas were prepared. The dried adult mosquito pools were crushed and homogenized with 200 μl of Leibovitz’s media (L-15) twice with a Teflon pestle homogenizer before centrifuging at 1000 rpm, 4°C for 10 minutes. The supernatant was aliquoted in tubes and stored at –80°C until further use.

### RNA extraction and DENV detection

The viral RNA from the homogenized mosquito supernatant was extracted using HiPurA pre-filled medium plates-T kit (HiMedia, Maharashtra, India) using a KingFisher Flex automated extraction system (Thermo Fisher Scientific, Waltham, USA). The mosquito pools were screened for DENV using a DENV real-time reverse transcriptase PCR kit (Helini Biomolecules, Chennai, India) in the Quant Studio 5 Real-time PCR System (Applied Biosystems, Waltham, USA) according to the manufacturer’s instructions. The kit contained pan-DENV-specific primers and probes for the quantification of DENV1-4 in the FAM channel. The target sequence 5’UTR is highly conserved across all DENV serotypes. The linear range of the assay kit ranged from 1 to 1×10^9^ copies/µl. Possible PCR inhibition and RNA purification efficiency were controlled using an internal amplification control in the HEX channel. In the RT-PCR assay, the cycle threshold (Ct) cut-off value range for DENV positivity was between 13 and 35. Any Ct value >35 was considered DENV-negative while a value <13 was diluted and the assay was repeated.

### Study design and participants

A community-based, cross-sectional, randomized cluster sampling was carried out to assess the seroprevalence of dengue in all 38 districts of Tamil Nadu, India. The study was approved by the Directorate of Public Health and Preventive Medicine, Government of Tamil Nadu and the Institutional Ethical Committee of the Madras Medical College (Approval No.:03092021). All individuals were aged >10 years and accented/consented to participate in the investigation. The individuals also included those with suspected or confirmed past dengue infection. From the 38 states, a total of 186 clusters were selected using stratified random sampling.

The size of the cluster was determined based on the population-to-size ratio and was considered as an adequate representation of the state. After identifying the cluster, the houses within the cluster were marked and numbered. During the study, a random household was selected and considered as the first household for the study and at least 30 to the left of the primary house were included in the study. The survey team collected all the identification details of the members including socio-demographic details from the selected household. From each household, one respondent was randomly identified for survey sampling using the Kish grid method. Participants were also given a unique ID for identification. At the time of sampling, the participants were enquired about dengue and COVID-19 status, vaccination status, and the type of SARS-CoV-2 vaccine administered.

### Clinical specimens

Considering a 76·9% dengue seroprevalence, a design effect of two, a confidence level of 95% and a precision value of 1·3, the required sample size was calculated as 20. Assuming one-third of the randomly assigned sample would become ineligible due to hemolysis during transportation and refusal to participate in the study, the final sample size was established as 30 per cluster. Two millilitres of venous blood was collected for serum separation before transporting to the District Public Health Laboratory for dengue IgM and IgG ELISA. The other aliquot was transported to the State Public Health Laboratory for SARS-CoV-2 IgG assay.

### Anti-DENV IgM and IgG

The extracted serum was tested for IgM as well as IgG antibodies using Panbio Dengue IgM capture ELISA (Abbot Diagnostics, South Korea). Cut-off values were determined as per the manufacturers’ instructions. Panbio Units (PU) were calculated as 10 times the value of sample absorbance divided by the cut-off value. A PU value >11 and <9 was considered positive and negative, respectively. Any value between 9 and 11 was considered equivocal, and was tested with the same assay and considered negative if the repeat test value was between 9 and 11. For anti-DENV IgG, a PU value of >22 and <18 were taken as positive and negative, respectively. Any value between 18 and 22 was considered equivocal, and was tested with the same assay and considered negative if the repeat test value was between 18 and 22.

### Anti-SARS-CoV-2 IgG

The serum samples were tested for SARS-CoV-2 IgG using a commercial anti-SARS-CoV-2 spike-specific quantitative IgG (VITROS S-IgG) assay (Ortho VITROS Immunodiagnostics, New Jersy, USA) as per manufacturers’ instructions. The assay kit detects anti-SARS-CoV-2 antibodies, and is FDA-approved under Emergency Use Authorization. The measuring range (or linearity) of the kit was 2-200 BAU/ml. However, based on the limit of quantitation, values ≥17·8 BAU/ml were considered reactive, and otherwise non-reactive.

### Statistical analysis

DENV seroprevalence was estimated using the IgM and IgG levels and corrected using a pre-assessed sensitivity and specificity of the same tests. The corrected prevalence was calculated using the formula: (apparent prevalence + specificity - 1)/(sensitivity + specificity - 1). Force of infection (FOI) was calculated for estimating the seroprevalence in each district, using the WHO-FOI calculator, which assumes a constant FOI over time. The relationship between total population, population density, and mosquito clusters positive for DENV and DENV seropositivity in clinical samples was evaluated using binary logistic regression. The factors associated with DENV seropositivity as well as anti-DENV IgM/IgG levels were evaluated using binary and linear logistic regressions, respectively. Statistical analyses were performed using PRISM, ver.5·02 (GraphPad, San Diego, CA). Binary and linear regression was performed using SPSS, ver.20 (IBM, Armonk, NY), Two-tailed P<0.05 was considered as significance, and P<0.05, <0.01, <0.001, were marked as *, ** and ***, respectively.

## Results

### DENV vector surveillance during 2017-24

To analyze the distribution of DENV-positive mosquitoes, the year-wise surveillance data of the State Public Health Laboratory, Chennai, and the Institute of Vector Control and Zoonoses, Hosur, Department of Public Health, Tamil Nadu between January 2016 and April 2024 were used for the comparison. The highest number of DENV-positive mosquito pools were observed during 2019, with 1440 of 3383 mosquito pools (42·6%). Though there was a two- to five-fold increase in the number of mosquito pools tested in subsequent years, there was a sudden decline during the SARS-CoV-2 pandemic with 8% positivity, and the decreasing trend in DENV-positivity until mid-2024. In succeeding years from 2020 until April 2024, the DENV-positivity remained 3-8% (**Figure 1**).

**Figure 1:**
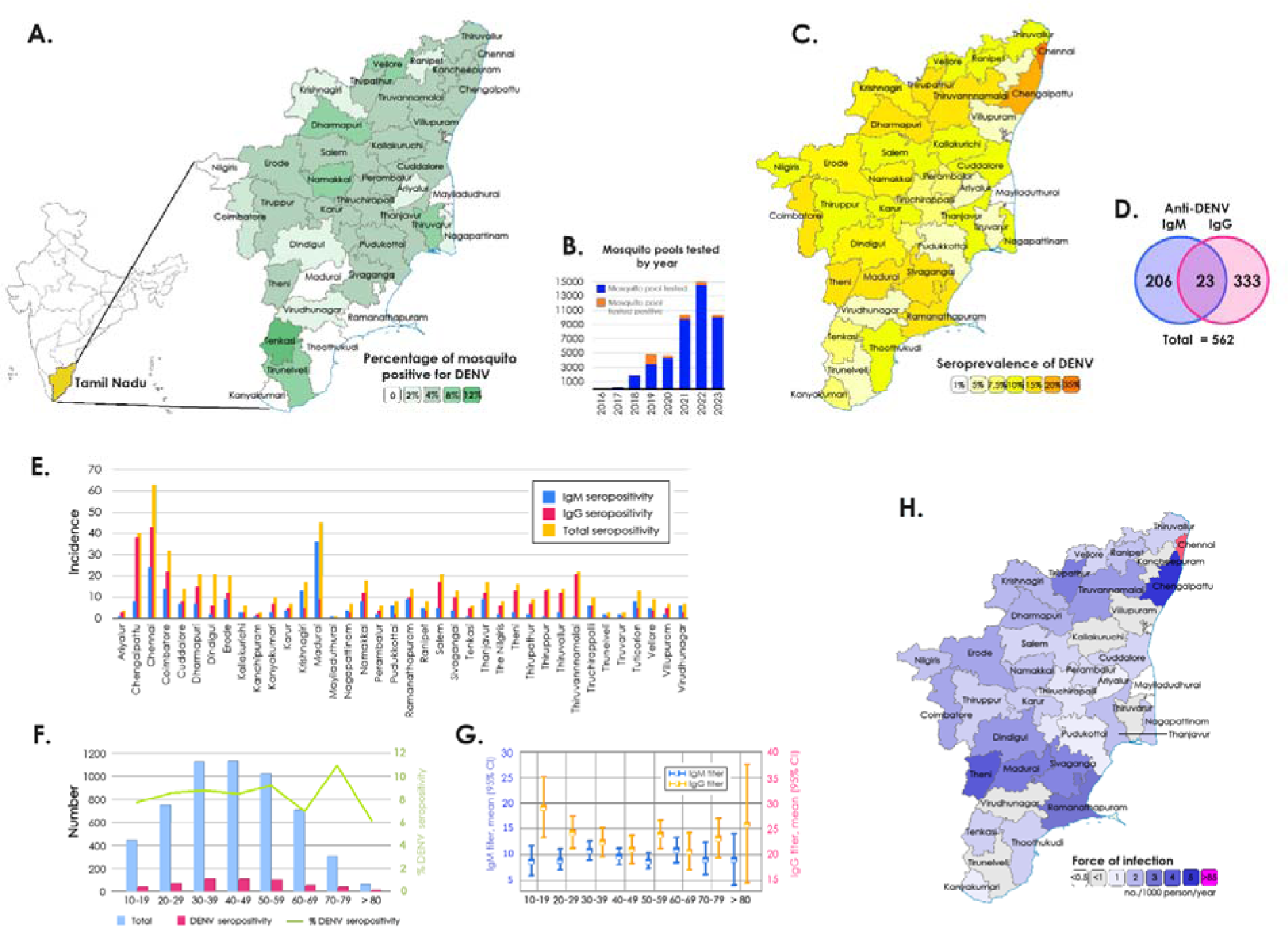
**A)** Spatial distribution of DENV-positive mosquito pools in Tamil Nadu, **B)** Mosquito pools tested DENV positive by year, **C)** Spatial distribution of seroprevalence of anti-DENV, **D)** Number of seropositivities for anti-DENV IgM and IgG, **E)** Distribution of DENV-seropositivity by districts, **F-G)** Distribution of DENV-seropositivity across different age groups **H)** Estimation of the force of infection (FOI) of DENV.

### DENV infestation rate in mosquito pools

Next, we analysed the concurrent seroprevalence of dengue and SARS-CoV-2 in 2021. We noticed a surge in global COVID-19 burden when mass vaccination programs were rolled-out by the Government of India. Of a total of 9764 Aedes mosquito pools tested, 387 (3·96%) tested positive for DENV (**Figure 1**). A decline in the distribution of DENV-infected Aedes mosquitoes was observed during the survey period compared to previous years. Of the 38 districts, Madurai recorded the highest number (n=644) of mosquito pools although the number of DENV-positive mosquito pools was highest in Tenkasi (11·1%) followed by Tirunelveli (8·07%) and Dharmapuri districts (7·89%). All the seven vector pools of the Nilgiris turned negative for DENV (**Figure 1**). The district-wise distribution of DENV in mosquito pools is presented in **Supplemental Table 1**.

### Anti-DENV IgM and IgG seroprevalence in December 2021

Of the 5577 serum samples collected from 186 clusters, 230 samples (4·12%) were positive for anti-DENV IgM whereas 360 (6·4%) were positive for anti-DENV IgG. The highest seroprevalence of anti-DENV IgG was reported in Chennai with 24% while Madurai and Chennai districts recorded the highest (13%) seroprevalence of anti-DENV IgM (**Figure 1**). The age of the recruited population ranged from 10-96 years with a median of 43·6 years (**Table 1**). A high anti-DENV IgM positivity was observed among patients with 30-39 years (n=51; 4·53%), 40-49 years (n=52; 4·59%) and 80-89 years (n=3; 4·84%) of age. Anti-DENV IgG positivity was higher among patients with 10-19 years (n=31; 7·01%), 20-29 years (n=54; 7·16%) and 70-79 years (n=28; 8·95%) of age. All individuals aged between 90 and 99 years (n=7) were negative for both DENV IgM and IgG. The IgM and IgG levels among male were 4·85% and 5·97%, whereas it was 3·57% and 6·81% among females, respectively. Anti-DENV IgM and IgG seroprevalence showed no significant difference among different age groups and between two genders. As the number of participants from the transgender community was low (n=4), no analyses could be performed. The association of patients’ domiciliary status (rural and urban) with total DENV seropositivity was highly significant. The seroprevalence of DENV in 38 districts of Tamil Nadu and the FOI in each district are listed in **Supplemental Tables 2a** and **2b**. Overall, the IgG seroprevalence and DENV-positivity in mosquito pools showed low (6·45% and 3·96%, respectively) during the study tenure.

**Table 1:** Socio-demographic, clinical and serological characteristics of the study participants.

|  |  |
| --- | --- |
| **Total number of participants, *n*** | 5577 |
| **Age, year; median (*IQR*)** | 43 (31 – 56) |
| **Gender, male; *n* (%)** | 2329 (41.8%) |
| **SARS-CoV-2 vaccination status, yes; *n* (%)** | 4654 (83.4%) |
| AZD1222; *n* (%) | 4249 (76.2%) |
| BBV152, *n* (%) | 395 (7.1%) |
| Others, *n* (%) | 10 (0.2%) |
| **History of SARS-CoV-2 infection; *n* (%)** | 166 (3%) |
| **Hospital admission, *n* (%)** | 77 (1.4%) |
| **SARS-CoV-2 IgG positivity, *n* (%)** | 4868 (87.3%) |
| **SARS-CoV-2 IgG titer, median (*IQR*)** | 200 (80.1 – 200) |
| **History DENV infection; *n* (%)** | 15 (0.3%) |
| **DENV IgM positivity; *n* (%)** | 229 (4.1%) |
| **DENV IgG positivity; *n* (%)** | 356 (6.4%) |
| **DENV IgM titer; median (*IQR*)** | 2.5 (1.53 – 4.10) |
| **DENV IgG titer; median (*IQR*)** | 5.10 (2.42 – 9.54) |

The association of seroprevalence (IgM/IgG/total) with DENV-positive mosquito pools was investigated using a simple linear regression model with a 95% CI of the slope (**Figure 2A-C**). The association was significantly correlating with IgM and total seropositivity, but not with IgG. The total DENV positivity was compared with DENV-positive mosquito clusters and DENV-FOI, which revealed no significance, which although was evident with DENV-seropositivity (**Figure 2D-F**). We also observed that DENV seroprevalence correlated significantly with factors such as total population, DENV-infested mosquito clusters and domiciliary status of participants (rural/urban) with increased odds (**Figure 2G**). The district-wise population-based seropositivity for SARS-CoV-2 IgG showed a high titre ranging from 78-97% with a mean titre of 167 IU/ml (**Figure 3A**). The number of samples positive for IgG in each district is listed in **Supplemental Table 3**.

**Figure 2:**
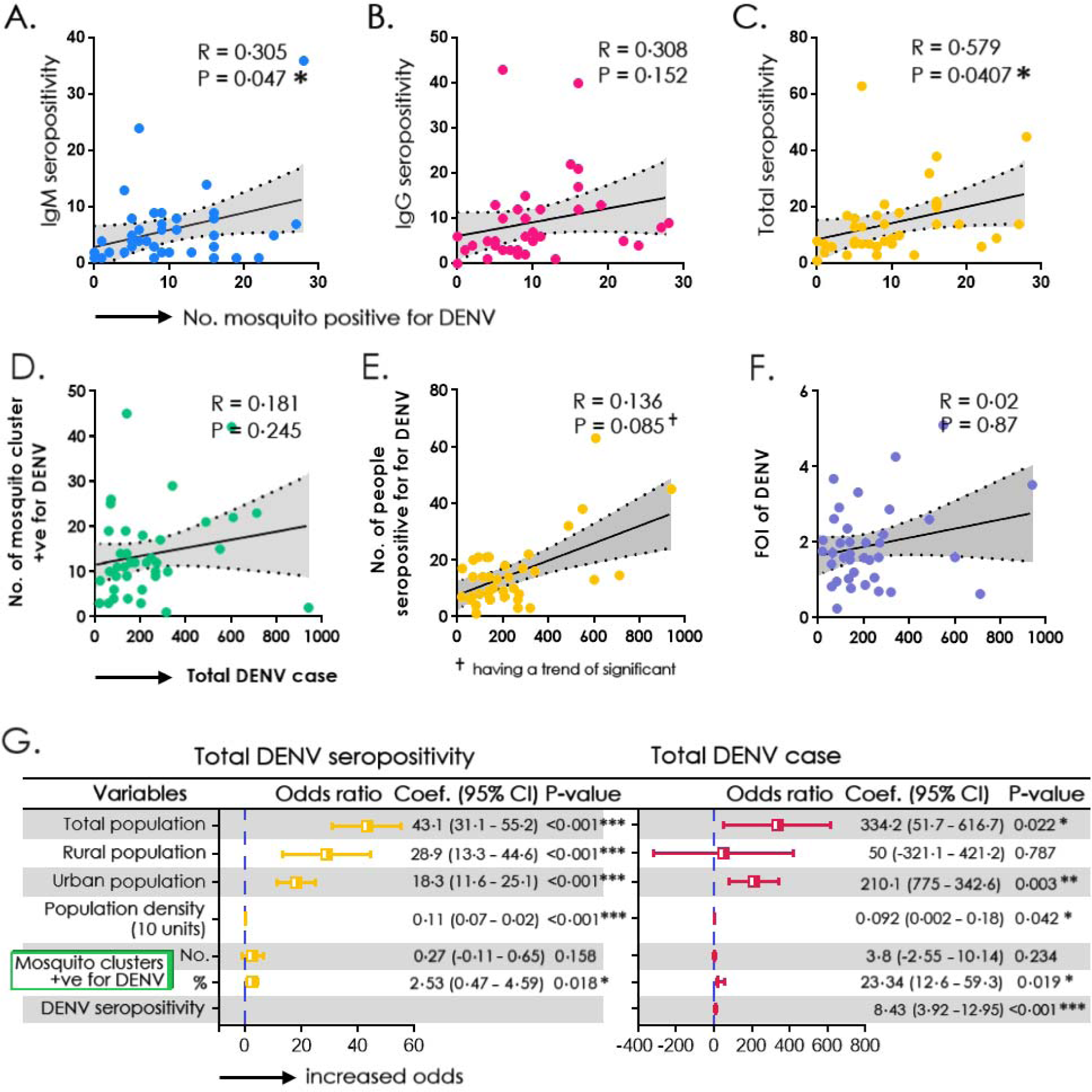
Correlation between a cluster of DENV-positive mosquito pools with **A)** DENV-IgM seropositivity, **B)** DENV-IgG seropositivity, **C)** Total DENV-seropositivity. Correlation between total anti-DENV IgM positive cases with **D)** Number of clusters of DENV-infested mosquito pools **E)** Number of individuals seropositive for DENV, and **F**) Force of infection of DENV, **G)** Simple binary regression model assessing the relationship between total population, population density and mosquito clusters positive for DENV with total (IgG and IgM) seropositivity and anti-DENV IgM positive cases.

**Figure 3:**
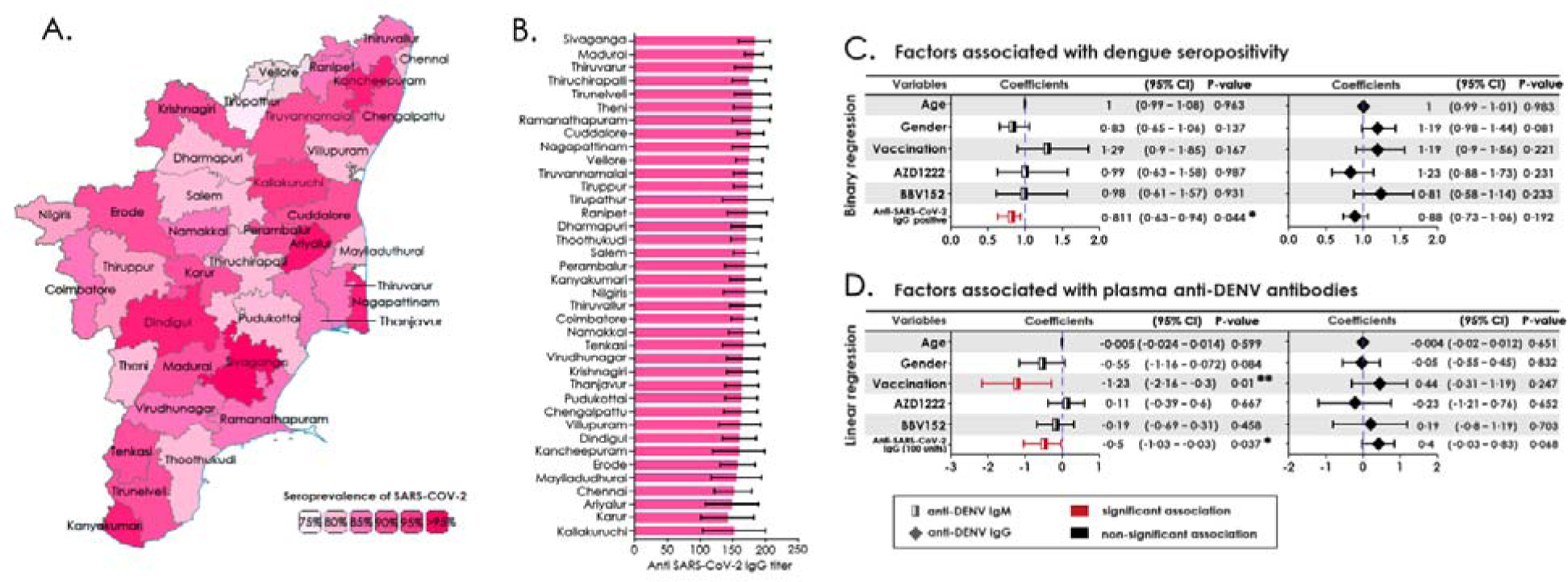
**A)** District-wise distribution of anti-SARS-CoV-2 seropositivity in Tamil Nadu. **B)** Average levels of anti-SARS-CoV-2 IgG titer **C)** Binary regression model assessing the factors associated with the anti-DENV IgM and IgG seropositivity, **D)** Linear regression model assessing the factors associated with the level of anti-DENV IgM and IgG.

Of the 5577 samples tested, 88·97% were reactive to SARS-CoV-2 IgG. Other factors including age, sex, vaccination status, type of vaccine administered and anti-SARS-CoV-2 antibodies were strongly associated with dengue seropositivity. While the levels of anti-SARS-CoV-2 correlated significantly with dengue seropositivity, vaccination status correlated similarly with anti-DENV IgM (**Figure 3C**). The association of two different types of SARS-CoV-2 vaccines, viz., BBV152 and AZD1222 with either anti-DENV IgM or IgG positivity did not reveal any significant difference (**Figure 3D**). The comparison of variables like total population, population density and number of DENV-positive mosquito clusters with DENV positivity is presented in **Supplemental Table 4a** and **4b**. The comparison of variables viz., age, gender, vaccination status and type of vaccine administered with DENV seropositivity is presented in **Supplemental Table 5a** and **5b**.

## Discussion

The burden of dengue fever and FOI poses considerable public health challenge to global health. However, the incidence is often underreported as most of the cases remain asymptomatic or misdiagnosed. The WHO data on global burden recorded the highest number of dengue cases (>6 million) and deaths (>7300) in 2023. DENV, being an arbovirus, has an ineludible link between human mobility, anthropogenic, and ecological factors^10^. Climate change and the spatio-temporal distribution of vectors due to *El Niño* cycle, urbanization, population density and human mobility patterns represent key risk factors driving viral transmission and incidence rates^21,22^. In countries like India, despite the implementation of systematic vector surveillance/control programs and access to specific diagnostic tools, increased incidence of dengue fever poses a significant socio-economic burden^23^. An effective and all-inclusive vector and disease control program must therefore include serological, molecular and entomological surveillance for real-time monitoring of DENV circulation.

Both DENV and SARS-CoV-2 are associated with a high risk of severe disease and mortality rate. SARS-CoV-2 evloved into a pandemic in 2020 and thus far recorded >775·52 million cases and 7.05 million deaths until 31 May 2024. After the first vaccine was rolled-out in mid-December 2020, 5·5 billion doses have been administered globally. At least 56% of the global population is vaccinated at least with a complete primary series of COVID-19 vaccines and 28% of the population is vaccinated with at least one booster dose (World Health Statistics, WHO; available at www.who.int/data; last accessed on 01 June 2024). Immunization with highly effective and safe vaccines that produced a high titre of nAb reduced the disease burden significantly. However, emerging infections due to the circulating variants of concern with increased transmissibility and severity still pose a serious threat to global health. The vaccine-derived nAbs did not offer cross-reactivity against the emerging new variants due to immune evasiveness influencing low transmission^24,25^. Serological cross-reactivity of anti-SARS-CoV-2 with dengue and zika viruses, especially in DENV endemic countries has been demonstrated previously^26^.

The two viruses, despite having different routes of entry into their host, have similar pathogenesis, overlapping clinical presentations posing diagnostic predicament and patient management. In addition, the antigenic similarities, heteroserotypic infection, and varying immunogenicity of the four DENV serotypes largely remain ambiguous. Hence, it is necessary to comprehensively analyze both the seroprevalence of DENV and SARS-CoV-2 at the community level to address the conundrum. In our study, the anti-DENV IgM and IgG titres were correlated with SARS-CoV-2 IgG in the community along with the virus screening in mosquitoes during December 2021. This was when both the cases and deaths declined upon vaccination in the timeline of the COVID-19 pandemic. The study was conducted across the state of Tamil Nadu, covering 38 administrative districts that had a population of 72 million. The state also recorded 2410 DENV cases in 2020; 6039 cases in 2021; 6430 cases in 2022 and 4148 cases in 2023 (as of September 2023) indicating its high endemicity for DENV.

The co-infection of DENV and SARS-CoV-2 further affects prognosis with increased mortality compared to either infection^27^. Convincing evidence of reduced dengue disease transmission was attributed to public health and social measures during the COVID-19 pandemic^10^. Reports of increased immature stages of Aedes mosquitoes due to COVID-19 lockdowns and subsequently interrupted larval control activities were expected to increase intra-household vector exposure and virus transmission^28^. A few countries reported increased dengue incidence during lockdowns^10,29^, but a decline in cases was observed in a vast majority of countries^17^.

India is one of the five highly endemic countries for dengue disease despite improved case management with a reduction in case-fatality rate to <0.5%. The Southeast Asian countries have witnessed a 46% upsurge in dengue cases between 2015 and 2019. A recent cross-sectional population-based serosurvey indicated 48·7% seroprevalence in India with the southern part of India which covered five states including Tamil Nadu recording the highest (77%). This indicated a high level of dengue transmission and geographical heterogeneity in the community during the pre-COVID-19 times^30^. A recent study reported a whopping 44·1% decrease in dengue across many dengue endemic regions, beginning March 2020 (2·2 million cases in 2020 versus 4·08 million in 2019)^10^. In Tamil Nadu, a downward trend was observed in dengue-positivity from 8527 cases in 2019 to 2410 in 2020 followed by 6039 cases in 2021 and 6430 in 2022. In 2023, we recorded 4524 cases until September 2023 followed by a sudden spike in dengue cases during the post-monsoon season (October to December) with a total of 10570 cases. This could be attributed to related changes in social activities before and after the pandemic, cross-reactive serological tests with SARS-CoV-2 or increased and improved testing capacity of global laboratories.

Several limitations encountered in previous studies were addressed by including both IgM and IgG assays, testing of large clusters, and proportionate number of mosquito pools in our study design. We showed a district-wise distribution of DENV-positive mosquito pools ranging from 1-7.3% with an average of 3.8%. Seroprevalence of IgM ranged from 0.6-13.3% with an average of 4.12% and IgG prevalence ranged from 1-23.9% with an average of 6.45%. The anti-SARS-CoV-2 IgG prevalence was high ranging from 78.3-97.8%. Interestingly, the anti-SARS-CoV-2 IgG titers correlated with dengue seropositivity indicating possible cross-protection, which however is unclear. Our data on vaccination status correlating with anti-DENV IgM levels needs further substantiation.

The low incidence of dengue during the COVID-19 pandemic due to misdiagnosis or underreporting cannot be ruled out. Hence, all the intense measures to curtail disease transmission should be ascertained. The canonical analysis of virus distribution in mosquitoes together with seroprevalence in the human population should be an integral part of public health measures to curb mosquito populations and pathogen transmission. Further, in addition to inclusion of both IgM and IgG seroprevalence, molecular typing could add details to the circulating dengue serotype and disease severity in the population.

The current study indicated the potential role of high titres of pre-existing anti-SARS-CoV-2 against DENV. However, the study suffered certain limitations that include the inability to demonstrate (1) a correlation of low titer of anti-SARS-CoV-2 with high anti-DENV seroprevalence in the population to further convince our findings; (2) cross-reactivity of the two viruses and viral interference by the immune cells in human cell lines; (3) deviations in patient’s clinical outcome; (4) DENV serotyping of the circulating viruses; (5) association of human mobility, public health, and social constraints. The inclusion of these factors will prove any association between the DENV infection and SARS-CoV-2 infection and if was a one-time phenomenon or observed at every emergence of SARS-CoV-2 variants across the globe.

In conclusion, dengue fever and SARS-CoV-2 continue to remain major global public health concerns, predominantly in the tropical world where dengue case incidence is exponentially increasing annually, and there is an ongoing geographical expansion of transmission areas and cocirculation of multiple DENV serotypes. It is of paramount importance to establish laboratory-based sentinel surveillance with coordinated entomological and molecular surveillance for early diagnosis, prevention, and control of arboviral infections.

## Data Availability

All data produced in the present study are available upon reasonable request to the corresponding authors.

## Contributors

STS, AS, SP, PK, RA, ABKC, SS, YKY, AM, ML, PB, SNB, VV, APD, EMS and SR designed the study and were responsible for conceptualization and data curation. STS, AARA, SC, YKY, PA, RPP, SS, AK, NG, MK, ML, VV, PB, APD, EMS and SR conducted the analysis and were responsible for methodology, formal analysis, validation, and visualization. STS, YKY, SS, ML, VV, EMS and SR wrote the first draft of the manuscript. All authors provided critical inputs and approved the final version of the manuscript for publication. All authors fulfil the criteria for authorship as per the ICMJE recommendations. All authors confirm that they have full access to all the data in the study and accept responsibility to submit it for publication.

## Data sharing

The data that support the findings of this study including deidentified participant data and specific datasets will be available from the corresponding author upon reasonable request by email. The data will be available beginning five months and ending three years after publication. Data requests can be sent to the corresponding author through email.

## Acknowledgements

The authors thank the contributions made by all the entomologists of the Department of Public Health across the state for their vector surveillance at the community level and referral of mosquito pools to SPHL, Chennai and IVC&Z, Hosur, India for the detection of DENV by RT-PCR assay.

