## Supplemental Table 1 for "Serosurveillance of dengue infection and correlation with mosquito pools for dengue virus positivity during the COVID-19 pandemic in Tamil Nadu, India – A state-wide cross-sectional cluster randomized community-based study"

### Supplemental Table 1: District-wise DENV positivity in mosquito pools in 2023

| S.No | District | No. of mosquito pools | | |
|---|---|---|---|---|
| | | Tested | DENV positivity, n (%) | |
| 1 | Ariyalur | 128 | 4 | 3·13 |
| 2 | Chengalpattu | 353 | 15 | 4·25 |
| 3 | Chennai | 302 | 22 | 7·28 |
| 4 | Coimbatore | 297 | 21 | 7·07 |
| 5 | Cuddalore | 460 | 12 | 2·61 |
| 6 | Dharmapuri | 217 | 11 | 5·07 |
| 7 | Dindigul | 425 | 9 | 2·12 |
| 8 | Erode | 427 | 26 | 6·09 |
| 9 | Kallakkurichi | 272 | 4 | 1·47 |
| 10 | Kancheepuram | 248 | 10 | 4·03 |
| 11 | Kanniyakumari | 284 | 13 | 4·58 |
| 12 | Karur | 267 | 3 | 1·12 |
| 13 | Krishnagiri | 626 | 17 | 2·72 |
| 14 | Madurai | 137 | 2 | 1·46 |
| 15 | Mayiladuthurai | 170 | 6 | 3·53 |
| 16 | Nagapattinam | 257 | 10 | 3·89 |
| 17 | Namakkal | 399 | 18 | 4·51 |
| 18 | Perambalur | 183 | 3 | 1·64 |
| 19 | Pudukkottai | 344 | 19 | 5·52 |
| 20 | Ramanathapuram | 448 | 24 | 5·36 |
| 21 | Ranipet | 246 | 11 | 4·47 |
| 22 | Salem | 834 | 43 | 5·16 |
| 23 | Sivaganga | 301 | 12 | 3·99 |
| 24 | Tenkasi | 434 | 10 | 2·30 |
| 25 | Thanjavur | 254 | 8 | 3·15 |
| 26 | The Nilgiris | 282 | 12 | 4·26 |
| 27 | Theni | 479 | 29 | 6·05 |
| 28 | Thiruvallur | 280 | 14 | 5·00 |
| 29 | Thiruvarur | 543 | 23 | 4·24 |
| 30 | Thoothukkudi | 582 | 41 | 7·04 |
| 31 | Tiruchirappalli | 338 | 12 | 3·55 |
| 32 | Tirunelveli | 329 | 9 | 2·74 |
| 33 | Tirupathur | 529 | 9 | 1·70 |
| 34 | Tiruppur | 673 | 14 | 2·08 |
| 35 | Tiruvannamalai | 31 | 1 | 3·23 |
| 36 | Vellore | 303 | 3 | 0·99 |
| 37 | Villupuram | 313 | 6 | 1·92 |
| 38 | Virudhunagar | 469 | 18 | 3·84 |
| **Total** | | **13464** | **524** | **3·76** |
