## Supplemental Table 2 for "Serosurveillance of dengue infection and correlation with mosquito pools for dengue virus positivity during the COVID-19 pandemic in Tamil Nadu, India – A state-wide cross-sectional cluster randomized community-based study"

### Supplemental Table 2: District-wise distribution of DENV seropositivity

| Sl. No. | District | Anti-DENV | | | | | Corrected | | | Force of infection (FOI) |
|---|---|---|---|---|---|---|---|---|---|---|
| | | IgM +ve | | IgG +ve | | Total Sero +ve | IgM +ve % | IgG +ve % | Total sero +ve % | |
| | | No. | % | No. | % | | | | | |
| 1 | Ariyalur | 1 | 1·11 | 3 | 3·33 | 4 | 0·216 | 2·33 | 3·60 | 0·000957 |
| 2 | Chengalpattu | 8 | 4·44 | 40 | 22·22 | 38 | 3·598 | 23·94 | 22·67 | 0·0051 |
| 3 | Chennai | 24 | 13·33 | 43 | 23·89 | 63 | 13·768 | 25·85 | 38·56 | 0·08689 |
| 4 | Coimbatore | 14 | 5·19 | 22 | 8·15 | 32 | 4·445 | 7·84 | 12·07 | 0·002601 |
| 5 | Cuddalore | 7 | 3·89 | 8 | 4·44 | 14 | 2·962 | 3·60 | 7·41 | 0·001528 |
| 6 | Dharmapuri | 7 | 3·89 | 15 | 8·33 | 21 | 2·962 | 8·05 | 11·86 | 0·002363 |
| 7 | Dindigul | 2 | 1·11 | 6 | 3·33 | 21 | 0·216 | 2·33 | 11·86 | 0·002919 |
| 8 | Erode | 9 | 5·00 | 12 | 6·67 | 20 | 4·233 | 6·14 | 11·23 | 0·002617 |
| 9 | Kallakkurichi | 3 | 3·33 | 3 | 3·33 | 6 | 2·326 | 2·33 | 6·14 | 0·000779 |
| 10 | Kancheepuram | 1 | 1·11 | 2 | 2·22 | 3 | 0·216 | 1·06 | 2·33 | 0·000678 |
| 11 | Kanniyakumari | 3 | 1·67 | 7 | 3·89 | 10 | 0·420 | 2·96 | 4·87 | 0·001058 |
| 12 | Karur | 4 | 4·44 | 5 | 5·56 | 7 | 3·598 | 4·87 | 7·41 | 0·00176 |
| 13 | Krishnagiri | 13 | 7·22 | 5 | 2·78 | 17 | 6·776 | 1·69 | 9·32 | 0·002205 |
| 14 | Madurai | 36 | 13·38 | 9 | 3·35 | 45 | 13·825 | 2·34 | 17·65 | 0·00352 |
| 15 | Mayiladuthurai | 1 | 1·11 | 0 | 0·00 | 1 | 0·216 | 1·49 | 0·22 | 0·000238 |
| 16 | Nagapattinam | 4 | 4·44 | 3 | 3·33 | 7 | 3·598 | 2·33 | 7·41 | 0·001723 |
| 17 | Namakkal | 8 | 4·44 | 12 | 6·67 | 18 | 3·598 | 6·14 | 9·95 | 0·001988 |
| 18 | Perambalur | 2 | 2·22 | 4 | 4·44 | 6 | 1·055 | 3·60 | 6·14 | 0·001423 |
| 19 | Pudukkottai | 6 | 3·33 | 2 | 1·11 | 8 | 2·326 | 0·22 | 3·60 | 0·001033 |
| 20 | Ramanathapuram | 9 | 10·00 | 10 | 11·11 | 14 | 9·954 | 11·23 | 16·31 | 0·003676 |
| 21 | Ranipet | 5 | 5·56 | 4 | 4·44 | 8 | 4·869 | 3·60 | 8·68 | 0·001981 |
| 22 | Salem | 5 | 1·85 | 17 | 6·30 | 21 | 0·631 | 5·72 | 7·41 | 0·001542 |
| 23 | Sivaganga | 4 | 4·44 | 10 | 11·11 | 13 | 3·598 | 11·23 | 15·04 | 0·003319 |
| 24 | Tenkasi | 1 | 1·11 | 5 | 5·56 | 6 | 0·216 | 4·87 | 6·14 | 0·001604 |
| 25 | Thanjavur | 9 | 5·00 | 12 | 6·67 | 17 | 4·233 | 6·14 | 9·32 | 0·002045 |
| 26 | The Nilgiris | 2 | 2·22 | 6 | 6·67 | 8 | 1·055 | 6·14 | 8·68 | 0·001981 |
| 27 | Theni | 3 | 3·33 | 13 | 14·44 | 16 | 2·326 | 15·04 | 18·85 | 0·004255 |
| 28 | Thiruvallur | 3 | 1·67 | 12 | 6·67 | 14 | 0·420 | 6·14 | 7·41 | 0·001603 |
| 29 | Thiruvarur | 2 | 2·22 | 1 | 1·11 | 3 | 1·055 | 0·22 | 2·33 | 0·000634 |
| 30 | Thoothukkudi | 8 | 4·49 | 5 | 2·81 | 13 | 3·655 | 1·73 | 6·87 | 0·001597 |
| 31 | Tiruchirappalli | 6 | 3·33 | 6 | 3·33 | 10 | 2·326 | 2·33 | 4·87 | 0·001203 |
| 32 | Tirunelveli | 2 | 2·22 | 1 | 1·11 | 3 | 1·055 | 0·22 | 2·33 | 0·000699 |
| 33 | Tirupathur | 2 | 3·33 | 7 | 11·67 | 9 | 2·326 | 11·86 | 15·68 | 0·001687 |
| 34 | Tiruppur | 1 | 0·56 | 13 | 7·22 | 14 | 0·852 | 6·78 | 7·41 | 0·002000 |
| 35 | Tiruvannamalai | 1 | 0·56 | 21 | 11·67 | 22 | 0·852 | 11·86 | 12·50 | 0·002865 |
| 36 | Vellore | 5 | 4·17 | 4 | 3·33 | 9 | 3·280 | 2·33 | 7·09 | 0·001591 |
| 37 | Villupuram | 2 | 1·11 | 5 | 2·78 | 7 | 0·216 | 1·69 | 2·96 | 0·000872 |
| 38 | Virudhunagar | 6 | 3·33 | 3 | 1·67 | 7 | 2·326 | 0·42 | 2·96 | 0·000826 |
| **Average** | | | | | | | **3·04** | **5·75** | **9·40** | **0·004141** |

### Supplemental Table 2 (continued)

| Sl. No. | District | Anti-DENV titer | | Force of infection | | DENV cases |
|---|---|---|---|---|---|---|
| | | IgM | IgG | FOI | no./1000 people/year | |
| 1 | Ariyalur | 2·27 | 2·4 | 0·000957 | 0·96 | 81 |
| 2 | Chengalpattu | 2·74 | 9·42 | 0·0051 | 5·10 | 550 |
| 3 | Chennai | 4·80 | 10·48 | 0·08689 | 86·89 | 608 |
| 4 | Coimbatore | 4·52 | 11·31 | 0·002601 | 2·60 | 488 |
| 5 | Cuddalore | 3·42 | 4·78 | 0·001528 | 1·53 | 226 |
| 6 | Dharmapuri | 3·83 | 9·62 | 0·002363 | 2·36 | 128 |
| 7 | Dindigul | 4·24 | 8·57 | 0·002919 | 2·92 | 92 |
| 8 | Erode | 5·38 | 10·56 | 0·002617 | 2·62 | 70 |
| 9 | Kallakkurichi | 4·67 | 3·25 | 0·000779 | 0·78 | 144 |
| 10 | Kancheepuram | 2·13 | 5·3 | 0·000678 | 0·68 | 320 |
| 11 | Kanniyakumari | 3·16 | 9·03 | 0·001058 | 1·06 | 247 |
| 12 | Karur | 4·09 | 7·05 | 0·00176 | 1·76 | 19 |
| 13 | Krishnagiri | 5·35 | 8·39 | 0·002205 | 2·20 | 287 |
| 14 | Madurai | 5·19 | 7·31 | 0·00352 | 3·52 | 940 |
| 15 | Mayiladuthurai | 3·23 | 2·87 | 0·000238 | 0·24 | 83 |
| 16 | Nagapattinam | 3·27 | 5·13 | 0·001723 | 1·72 | 63 |
| 17 | Namakkal | 3·46 | 7·59 | 0·001988 | 1·99 | 209 |
| 18 | Perambalur | 2·27 | 6·28 | 0·001423 | 1·42 | 59 |
| 19 | Pudukkottai | 3·71 | 2·66 | 0·001033 | 1·03 | 134 |
| 20 | Ramanathapuram | 5·32 | 9·32 | 0·003676 | 3·68 | 68 |
| 21 | Ranipet | 3·57 | 5·4 | 0·001981 | 1·98 | 97 |
| 22 | Salem | 3·35 | 9·23 | 0·001542 | 1·54 | 139 |
| 23 | Sivaganga | 3·64 | 8·97 | 0·003319 | 3·32 | 175 |
| 24 | Tenkasi | 2·22 | 8·85 | 0·001604 | 1·60 | 266 |
| 25 | Thanjavur | 5·61 | 8·01 | 0·002045 | 2·05 | 22 |
| 26 | The Nilgiris | 2·82 | 5·44 | 0·001981 | 1·98 | 271 |
| 27 | Theni | 4·05 | 12·12 | 0·004255 | 4·26 | 340 |
| 28 | Thiruvallur | 3·13 | 2·45 | 0·001603 | 1·20 | 111 |
| 29 | Thiruvarur | 2·45 | 4·04 | 0·000634 | 1·60 | 712 |
| 30 | Thoothukkudi | 4·4 | 6·87 | 0·001597 | 0·63 | 601 |
| 31 | Tiruchirappalli | 2·85 | 6·6 | 0·001203 | 1·60 | 157 |
| 32 | Tirunelveli | 2·92 | 6·47 | 0·000699 | 0·70 | 266 |
| 33 | Tirupathur | 4·52 | 12·64 | 0·001687 | 3·46 | 140 |
| 34 | Tiruppur | 2·18 | 11·16 | 0·002000 | 1·69 | 139 |
| 35 | Tiruvannamalai | 2·76 | 10·11 | 0·002865 | 2·87 | 313 |
| 36 | Vellore | 3·12 | 5·67 | 0·001591 | 1·59 | 202 |
| 37 | Villupuram | 1·27 | 4·68 | 0·000872 | 0·87 | 207 |
| 38 | Virudhunagar | 3·92 | 7·44 | 0·000826 | 0·83 | 60 |
| | **Average** | **2·94** | **8·1** | **0·004141** | **4·18** | |
| **Total** | | | | | | **9034** |
