## Supplemental Table 3 for "Serosurveillance of dengue infection and correlation with mosquito pools for dengue virus positivity during the COVID-19 pandemic in Tamil Nadu, India – A state-wide cross-sectional cluster randomized community-based study"

### Supplemental Table 3: District-wise distribution of SARS-CoV-2 seropositivity

| Sl. No. | District | Samples (n) | Anti-SARS-CoV-2 IgG | | | |
|---|---|---|---|---|---|---|
| | | | No. IgG +ve | % IgG +ve | Mean | 95% CI |
| 1 | Ariyalur | 90 | 87 | 96·67 | 149·03 | (132·87 - 165·19) |
| 2 | Chengalpattu | 180 | 166 | 92·22 | 162·13 | (151·87 - 172·39) |
| 3 | Chennai | 180 | 154 | 85·56 | 151·57 | (140·53 - 162·61) |
| 4 | Coimbatore | 270 | 236 | 87·41 | 166·7 | (158·94 - 174·46) |
| 5 | Cuddalore | 180 | 165 | 91·67 | 176·79 | (168·90 - 184·86) |
| 6 | Dharmapuri | 180 | 153 | 85·00 | 170·46 | (161·34 - 179·58) |
| 7 | Dindigul | 180 | 172 | 95·56 | 160·17 | (149·89 - 170·45) |
| 8 | Erode | 180 | 165 | 91·67 | 157·57 | (146·86 - 168·28) |
| 9 | Kallakuruchi | 90 | 84 | 93·33 | 140·72 | (140·91 - 174·23) |
| 10 | Kancheepuram | 90 | 86 | 95·56 | 159·6 | (143·76 - 175·44) |
| 11 | Kanyakumari | 180 | 172 | 95·56 | 168·62 | (159·26 - 177·98) |
| 12 | Karur | 90 | 84 | 93·33 | 141·56 | (125·26 - 157·86) |
| 13 | Krishnagiri | 180 | 164 | 91·11 | 164·5 | (155·22 - 173·78) |
| 14 | Madurai | 269 | 247 | 91·82 | 182·24 | (176·61 - 187·87) |
| 15 | Mayiladudhurai | 90 | 77 | 85·56 | 155·31 | (139·86 - 170·76) |
| 16 | Nagapattinam | 90 | 86 | 95·56 | 176·57 | (165·83 - 187·31) |
| 17 | Namakkal | 180 | 158 | 87·78 | 166·18 | (156·98 - 175·38) |
| 18 | Perambalur | 90 | 84 | 93·33 | 168·86 | (156·31 - 181·41) |
| 19 | Pudukottai | 180 | 152 | 84·44 | 163·18 | (153·34 - 173·03) |
| 20 | Ramanathapuram | 90 | 79 | 87·78 | 178·11 | (166·73 - 189·49) |
| 21 | Ranipet | 90 | 80 | 88·89 | 171·78 | (159·69 - 183·87) |
| 22 | Salem | 270 | 228 | 84·44 | 169·58 | (162·09 - 177·07) |
| 23 | Sivaganga | 90 | 88 | 97·78 | 182·77 | (173·08 - 192·46) |
| 24 | Tenkasi | 90 | 84 | 93·33 | 166·17 | (153·28 - 179·06) |
| 25 | Thanjavur | 180 | 158 | 87·78 | 163·67 | (153·56 - 173·78) |
| 26 | The Nilgiris | 90 | 78 | 86·67 | 168·31 | (155·37 - 181·26) |
| 27 | Theni | 90 | 77 | 85·56 | 179·8 | (168·43 - 191·17) |
| 28 | Thiruchirapalli | 180 | 145 | 80·56 | 180·64 | (162·81 - 180·31) |
| 29 | Thiruvallur | 180 | 160 | 88·89 | 168·2 | (158·86 - 177·54) |
| 30 | Thiruvarur | 90 | 80 | 88·89 | 180·64 | (169·47 - 191·81) |
| 31 | Thoothukudi | 178 | 149 | 83·71 | 170·26 | (161·05 - 179·47) |
| 32 | Tirunelveli | 90 | 83 | 92·22 | 180·04 | (169·24 - 190·84) |
| 33 | Tirupathur | 60 | 47 | 78·33 | 172·38 | (157·09 - 187·67) |
| 34 | Tiruppur | 180 | 155 | 86·11 | 172·68 | (164·01 - 181·35) |
| 35 | Tiruvannamalai | 180 | 167 | 92·78 | 172·68 | (164 - 181·35) |
| 36 | Vellore | 120 | 97 | 80·83 | 174·64 | (166·53 - 182·75) |
| 37 | Villupuram | 180 | 155 | 86·11 | 160·57 | (147·75 - 173·39) |
| 38 | Virudhunagar | 180 | 160 | 88·89 | 165·07 | (155·26 - 174·88) |
| **Total** | | | **4962** | | **167·36** | |
