## Supplemental Table 4 for "Serosurveillance of dengue infection and correlation with mosquito pools for dengue virus positivity during the COVID-19 pandemic in Tamil Nadu, India – A state-wide cross-sectional cluster randomized community-based study"

**Supplemental Table 4a: Mosquito pools positive for DENV and DENV seropositivity**

| Linear Regression | | | | |
|---|---|---|---|---|
| **Outcome** | **Total DENV seropositivity** | | | |
| **Variable** | **Media** | **95% CI** | | **P value** |
| Total population | 43·13 | 31·11 | 55·16 | <0·001*** |
| Rural | 28·96 | 13·28 | 44·64 | <0·001*** |
| Urban | 18·34 | 11·55 | 25·13 | <0·001*** |
| Population density (10 units) | 0·11 | 0·07 | 0·16 | <0·001*** |
| No. of mosquito clusters positive for DENV | 2·7 | -1·09 | 6·5 | 0·158 |
| % of mosquito clusters positive for DENV | 2·53 | 0·47 | 4·59 | 0·018* |

**Supplemental Table 4b: Mosquito positive for DENV and DENV positive case**

| Linear Regression | | | | |
|---|---|---|---|---|
| **Outcome** | **Total DENV positive cases** | | | |
| **Variable** | **Media** | **95% CI** | | **P value** |
| Total population | 334·2 | 51·66 | 616·74 | 0·022* |
| Rural | 50·04 | -321·1 | 421·21 | 0·787 |
| Urban | 210·07 | 77·54 | 342·6 | 0·003 |
| Population density (10 units) | 0·02 | 0·02 | 0·02 | <0·01** |
| No. of mosquito clusters positive for DENV | 3·8 | -2·55 | 10·14 | 0·234 |
| % of mosquito clusters positive for DENV | 23·34 | 12·6 | 59·28 | 0·02* |
