## Supplemental Table 5 for "Serosurveillance of dengue infection and correlation with mosquito pools for dengue virus positivity during the COVID-19 pandemic in Tamil Nadu, India – A state-wide cross-sectional cluster randomized community-based study"

### Supplemental Table 5a: Factors associated with DENV seropositivity

| Binary Regression | | | | | | | | |
|---|---|---|---|---|---|---|---|---|
| **Outcome** | **Anti-DENV IgM positivity** | | | | **Anti-DENV IgG positivity** | | | |
| **Variable** | **Media** | **95% CI** | | **P value** | **Media** | **95% CI** | | **P value** |
| Age | 1 | 0·993 | 1·008 | 0·963 | 1 | 0·994 | 1·006 | 0·983 |
| Gender | 0·832 | 0·653 | 1·06 | 0·137 | 1·187 | 0·979 | 1·44 | 0·081 |
| Vaccination status | 1·291 | 0·899 | 1·854 | 0·167 | 1·185 | 0·903 | 1·556 | 0·221 |
| AZD1222 | 0·996 | 0·629 | 1·578 | 0·987 | 0·814 | 0·581 | 1·141 | 0·233 |
| BBV152 | 0·979 | 0·612 | 1·567 | 0·931 | 1·232 | 0·877 | 1·732 | 0·23 |
| Anti-SARS-CoV-2 IgG (100 units) | 0·811 | 0·634 | 0·936 | 0·044* | 0·884 | 0·734 | 1·064 | 0·192 |

### Supplemental Table 5b: Factors associated with DENV IgM and IgG levels

| Linear Regression | | | | | | | | |
|---|---|---|---|---|---|---|---|---|
| **Outcome** | **Anti-DENV IgM positivity** | | | | **Anti-DENV IgG positivity** | | | |
| **Variable** | **Media** | **95% CI** | | **P value** | **Media** | **95% CI** | | **P value** |
| Age | -0·005 | -0·024 | 0·014 | 0·599 | -0·004 | -0·019 | 0·012 | 0·651 |
| Gender | -0·545 | -1·163 | 0·072 | 0·084 | -0·054 | -0·553 | 0·445 | 0·832 |
| Vaccination status | -1·227 | -2·156 | -0·298 | 0·01** | 0·443 | -0·307 | 1·194 | 0·247 |
| AZD1222 | 0·108 | -0·386 | 0·603 | 0·667 | -0·226 | -1·21 | 0·757 | 0·652 |
| BBV152 | -0·19 | -0·691 | 0·311 | 0·458 | 0·194 | -0·803 | 1·191 | 0·703 |
| Anti-SARS-CoV-2 IgG (100 units) | -0·501 | -1·03 | -0·028 | 0·037* | 0·398 | -0·029 | 0·825 | 0·068 |
